# Anti-neuronal IgG4 autoimmune diseases and IgG4-related diseases may not be part of the same spectrum: a comparative study

**DOI:** 10.1101/2021.09.30.21264258

**Authors:** Verena Endmayr, Cansu Tunc, Lara Ergin, Anna de Rosa, Rosa Weng, Lukas Wagner, Thin-Yau Yu, Andreas Fichtenbaum, Thomas Perkmann, Helmuth Haslacher, Nicolas Kozakowski, Carmen Schwaiger, Gerda Ricken, Simon Hametner, Lívia Almeida Dutra, Christian Lechner, Désirée de Simoni, Kai-Nicolas Poppert, Georg Johannes Müller, Susanne Pirker, Walter Pirker, Aleksandra Angelovski, Matus Valach, Michelangelo Maestri, Melania Guida, Roberta Ricciardi, Florian Frommlet, Daniela Sieghart, Miklos Pinter, Romana Höftberger, Inga Koneczny

**Affiliations:** Division of Neuropathology and Neurochemistry, Department of Neurology, Medical University of Vienna, Austria; Department of Clinical and Experimental Medicine, Neurology Unit, University of Pisa, Via Paradisa 2, 56124 Pisa, Italy; Department of Neurology, Medical University of Vienna, Austria; Department of Laboratory Medicine, Medical University of Vienna, Austria; Department of Pathology, Medical University of Vienna, Austria; Hospital Israelita Albert Einstein, Department of Neurology and Neurosurgery, São Paulo, Brazil; Pediatric Neurology, Department of Pediatric and Adolescent Medicine, Medical University of Innsbruck, Austria; Department of Neurology, University Hospital St. Poelten, St. Poelten, Austria; Department of Neurology, Christian Doppler University Hospital, Paracelsus Medical University, Salzburg, Austria; Department of Neurology and Karl Landsteiner Institute for Neuroimmunological and Neurodegenerative Disorders, Klinik Donaustadt, Vienna, Austria; Department of Neurology, Klinik Hietzing, Vienna, Austria; Department of Neurology, Klinik Ottakring, Vienna, Austria; Department of Pathology, Klinik Landstrasse, Vienna, Austria; Center for Medical Statistics, Informatics and Intelligent Systems, Section for Medical Statistics, Medical University of Vienna, Austria; Division of Rheumatology, Department of Internal Medicine III, Medical University of Vienna, Austria; Wiener Privatklinik – Health Center, Lazarettgasse 25, 1090 Vienna, Austria

**Keywords:** IgG4-related diseases, IgG4 autoimmune diseases, MuSK myasthenia gravis, CIDP, LGI1, Caspr2

## Abstract

**Background:** IgG4 is associated with two emerging groups of rare diseases: 1) IgG4 autoimmune diseases (IgG4-AID) and 2) IgG4-related diseases (IgG4-RLD). Anti-neuronal IgG4-AID include MuSK myasthenia gravis, LGI1- and Caspr2-encephalitis and autoimmune nodo-/paranodopathies (CNTN1 or NF155 antibodies). IgG4-RLD is a multiorgan disease hallmarked by tissue-destructive fibrotic lesions with lymphocyte and IgG4 plasma cell infiltrates and increased serum IgG4 concentrations. It is unclear, whether IgG4-AID and IgG4-RLD share relevant clinical and immunopathological features.

**Methods:** We collected and analysed serological, clinical, and histopathological data in 50 patients with anti-neuronal IgG4-AID and 16 patients with IgG4-RLD.

**Results:** A significantly higher proportion of IgG4-RLD patients had serum IgG4 elevation when compared to IgG4-AID patients (50% vs. 16%, *p* = .015). Moreover, those IgG4-AID patients with elevated IgG4 did not meet the diagnostic criteria of IgG4-RLD, and their autoantibody titres did not correlate with their serum IgG4 concentrations. In addition, patients with IgG4-RLD were negative for anti-neuronal/neuromuscular autoantibodies and among these patients, men showed a significantly higher propensity for IgG4 elevation, when compared to women (*p* = .041). Last, a kidney biopsy from a patient with autoimmune paranodopathy due to CNTN1/Caspr1-complex IgG4 autoantibodies and concomitant nephrotic syndrome did not show fibrosis or IgG4^+^ plasma cells, which are diagnostic hallmarks of IgG4-RLD.

**Conclusion:** Our observations suggest that anti-neuronal IgG4-AID and IgG4-RLD are most likely distinct disease entities.

## Introduction

In the last decade, two new groups of rare diseases emerged that are associated with the IgG4 subclass: 1) IgG4 autoimmune diseases (IgG4-AID), first appreciated as a distinct subgroup of autoimmune diseases in 2015 (1) and 2) IgG4-related diseases (IgG4-RLD), first described systematically in 2012 (2). Anti-neuronal IgG4-AID, which comprise the largest subgroup of IgG4-AID (3), include muscle-specific kinase (MuSK) myasthenia gravis (MG), leucine-rich glioma inactivated protein-1 (LGI1)- and contactin-associated protein-like 2 (Caspr2)-encephalitis and autoimmune nodo-/paranodopathies with autoantibodies against contactin 1 (CNTN1) or neurofascin-155 (NF155) (3). The diagnosis of IgG4-AID in patients presenting with disease-specific clinical symptoms (e.g. fatigable skeletal muscle weakness in MuSK-MG) is based on the detection of antigen-specific autoantibodies. IgG4-RLD is a multiorgan disease, and diagnostic criteria include organ enlargement, the presence of tissue-destructive fibrotic lesions with a storiform pattern, obliterative phlebitis, dense lymphocyte and IgG4^+^ plasma cell infiltrates and increased serum IgG4 concentrations (2, 4).

Patients with IgG4-RLD and concomitant IgG4-AID were reported in two single case reports (5, 6). They may co-occur by chance, as each of these disease groups is very rare (prevalence: <0.0001 – 5/10,000 in IgG4-AID, <0.01 – 8/10,000 in IgG4-RLD^1^ (3)), and the question arose whether these diseases may be related (5). We addressed this question by comparing serological, clinical, and histopathological findings in 50 patients with anti-neuronal IgG4-AID and 16 patients with IgG4-RLD to find out whether there are indications for an overlap between these diseases.

## Materials and Methods

### Patients

Sera of 50 patients (17 female, 33 male) with a clinical diagnosis of anti-neuronal IgG4 autoimmune disease (autoimmune encephalitis associated with LGI1 (n=15) or Caspr2 (n=9) autoantibodies, chronic inflammatory demyelinating polyneuropathy (CIDP) associated with anti-NF155 (n=2) or pan anti-NF155/140/186 (pan-NF, n=1), anti-CNTN1/Caspr1-complex (n=2), anti-CNTN1 (n=5) or anti-Caspr1 (n=1) autoantibodies and MuSK myasthenia gravis (n=15)) taken at the time of diagnosis or at the earliest time point available where a clear autoantibody titre was present, and sera of 53 patients with suspected neurometabolic disease (in which IgG concentrations are considered to be unaffected by disease, 30 female, 23 male) and from 13 healthy controls (eight female, five male) were selected. These were derived from archival blood samples that were sent for diagnostic purposes and stored at the biobank of the Division of Neuropathology and Neurochemistry, Department of Neurology, Medical University of Vienna, Austria (EK1123-2015). Archival nephelometry serum samples from the biobank of the Division of Rheumatology, Department of Internal Medicine III, Medical University of Vienna, Austria from 16 patients with IgG4-related diseases were analysed retrospectively (EK559/2005). The samples were processed and stored according to standard operating procedures at the Medical University of Vienna biobank in an ISO 9001-certified environment (7).

The study was approved by the Institutional Review Boards of the Medical University of Vienna, Austria (EK 1442/2017).

### Cell-based assay (CBA)

Briefly, live human embryonic kidney cells (HEK293T) were incubated with serum diluted 1:40 in medium (CNTN1/Caspr1, CNTN1, LGI1) or medium supplemented with 1% bovine serum albumin (BSA) (MuSK) for 30 (CNTN1/Caspr1, CNTN1, LGI1) or 60 minutes (min) (MuSK) at 37°C. Afterwards, cells were fixed with 4% cold paraformaldehyde (PFA; Alfa Aesar) for 10 min, permeabilized with 0.3% Triton X-100 (Merck) for 5 min (CNTN1/Caspr1, CNTN1 and LGI1 only) and incubated with a commercial antibody (anti-CNTN1, rabbit polyclonal, Sigma #HPA070467; anti-ADAM23, rabbit polyclonal, Abcam #ab28304 - both diluted in 1% BSA) for 60 min at room temperature (RT). Bound antibodies were detected with fluorescent-conjugated Alexa Fluor^®^ secondary antibodies against human (AF594) and rabbit IgGs (AF488) (both 1:750; diluted in PBS (CNTN1/Caspr1, CNTN1, LGI1) or culture medium supplemented with 1% BSA (MuSK) for 30 min (CNTN1/Caspr1, CNTN1, LGI1) or 45 min (MuSK) at RT in the dark). For nuclear staining, 4’,6-diamidino-2-phenylindole (DAPI) was used. The fluorescence was analysed with an OLYMPUS BX63 fluorescence microscope.

### Fixed cell-based assay

The cells were fixed with 4% cold PFA for 10 min, permeabilized with 0.3% Triton X-100 for 5 min and blocked for 1.5 hours with 1% BSA. Next, the cells were incubated with serum diluted 1:40 in PBS with 1% BSA over night at 4°C. The antigens were labeled using commercial antibodies (anti-Caspr2, rabbit polyclonal, Abcam #ab33994; anti-c-myc, mouse monoclonal, Roche #11667149001 – both diluted in 1% BSA) for 30 min at RT, and antibody binding was detected by incubation with anti-human (AF594) and mouse/rabbit IgGs (AF488) (both diluted 1:750 in PBS). Nuclei were stained with DAPI. The fluorescence was analysed with an OLYMPUS BX63 fluorescence microscope.

### Neuropathology

Immunohistochemical stainings were conducted as described in (8).

### Nephelometry

Human total serum IgG and subclass IgG4 concentrations were determined using particle-enhanced immune nephelometry with the BN II System (BN II Nephelometer, Siemens). The internal reference values for IgG4 were 0.03 - 2.01 g/L, and for total IgG 7-16 g/L. The published upper threshold for IgG4 in IgG4-RLD is 1.35 g/L. Serum concentrations ≥1.35 g/L were considered as elevated.

### Statistical analysis

Due to the heteroscedastic distribution of the data and small sample size of some groups, statistical analysis with ANOVA to compare serum IgG4 concentrations between the different disease groups was considered as inappropriate. Instead, we report the mean and 95% confidence interval for each group. Nonparametric Spearman correlation was used to analyse the relationship between total IgG and IgG4 concentrations per group and between autoantibody titres and serum IgG4 concentrations. In order to test gender- and disease-dependent proportions of IgG4 elevation in patients, Fisher’s exact tests were applied. Statistical analysis was conducted using GraphPad Prism 9 and IBM SPSS Version 27.

## Results

### Patients

Anti-neuronal/neuromuscular autoantibodies were identified in all 50 patients with anti-neuronal IgG4 autoimmune disease by cell-based assays (Figure 1). Clinical and epidemiological data of the study cohort is summarized in Supplementary Table 1.

**Figure 1:**
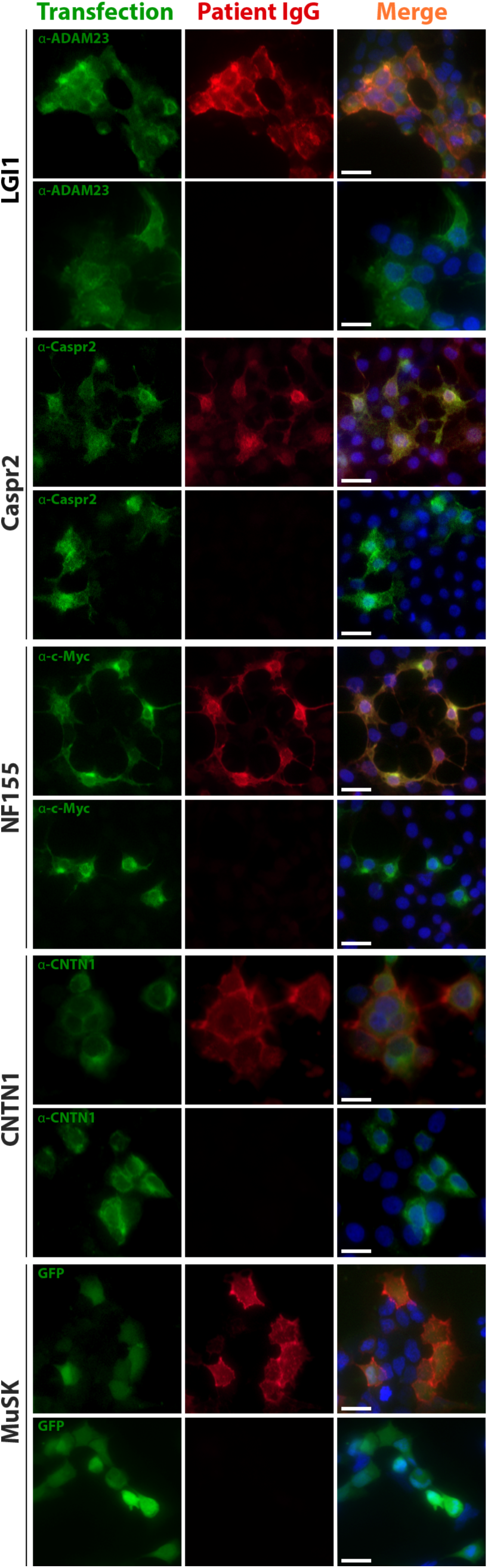
Anti-neuronal/neuromuscular autoantibodies were detected using cell-based assays. Example images of sera positive and negative for LGI1, Caspr2, NF155, CNTN1 and MuSK autoantibodies are shown. Green fluorescence indicates antigen expression by either counterstaining with commercial antibodies and secondary antibodies conjugated to AF488 or GFP co-expressed after an IRES site on the plasmid coding for MuSK. Red fluorescence indicates patient autoantibodies detected by anti-human IgG conjugated to AF594. Blue fluorescence corresponds to nuclear staining with DAPI. Scale bar = 20 µm. ADAM23 = disintegrin and metalloproteinase domain-containing protein 23; Caspr2 = contactin-associated protein-like 2; CNTN1 = contactin 1; GFP = green fluorescent protein; IgG = immunoglobulin type G; LGI1 = leucine-rich glioma inactivated protein- 1; MuSK = muscle-specific kinase; NF155 = neurofascin 155.

### Normal serum IgG4 concentrations in the majority of patients with IgG4-AID

Elevated serum IgG4 concentrations are frequently observed in patients with IgG4-RLD, but it is unknown if this is also the case in patients with anti-neuronal IgG4-AID. Total serum IgG and IgG4 concentrations were measured using nephelometry, and sera with concentrations reaching the published cut-off value of ≥1.35g/L IgG4 (9) were considered as elevated (Table 1). As expected, half of the patients with IgG4-RLD (8/16, 50%) had elevated serum IgG4 concentrations, while the majority of healthy (84.62%) and neurometabolic controls (92.45%) had normal serum IgG4 levels. Conversely, the majority of patients with IgG4-AID (84%) had normal serum IgG4 levels. When using a Fisher’s exact test, IgG4-RLD patients were significantly more likely to display elevated IgG4, compared to patients with IgG4-AID (likelihood ratio: 6.962, *p* = .015).

**Table 1:**
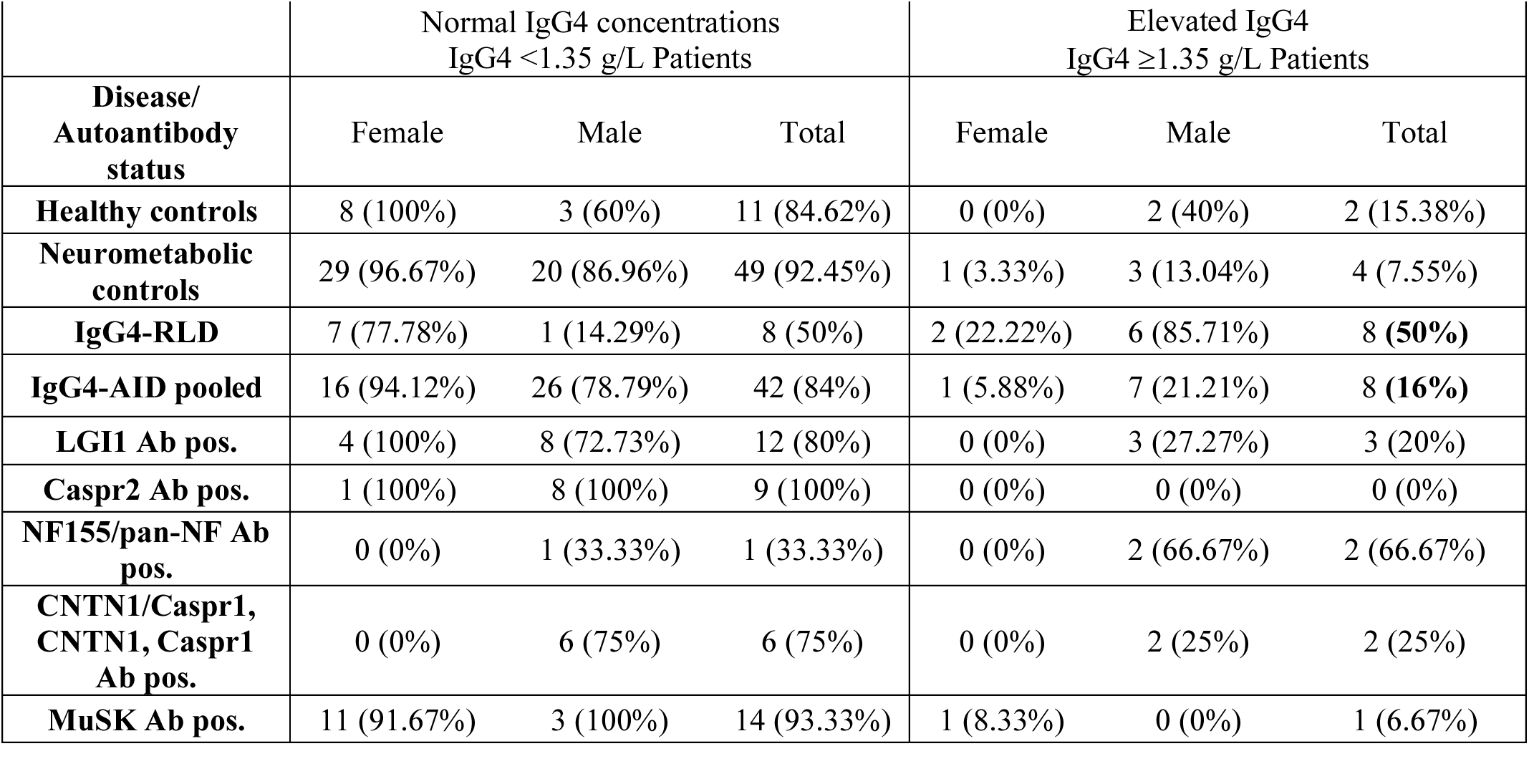
Number of patients with normal and elevated serum IgG4 concentrations in IgG4-AID and IgG4-RLD. Values indicate number of patients (female, male and total) per group (normal and elevated IgG4), percentages indicate percent of females, males or total of both groups (normal and elevated IgG4). Ab = antibody; Caspr1 = contactin-associated protein-like 1; Caspr2 = contactin-associated protein-like 2; CNTN1 = contactin 1; IgG4 = immunoglobulin type G subclass 4; IgG4-AID = IgG4 autoimmune disease; IgG4-RLD = IgG4-related disease; LGI1 = leucine-rich glioma inactivated protein-1; MuSK = muscle-specific kinase; NF155 = neurofascin 155; pan-NF Ab pos. = positive for pan neurofascin antibodies; pos. = positive.

The highest absolute serum IgG4 concentrations (Figure 2A) were observed in patients with IgG4-RLD (up to 17.1 g/L). Eight patients with LGI1, CNTN1/Caspr1-complex, NF155 or pan-NF and MuSK autoantibodies had elevated serum IgG4 concentrations but these were in a similar range as in six of the healthy and neurometabolic controls.

**Figure 2:**
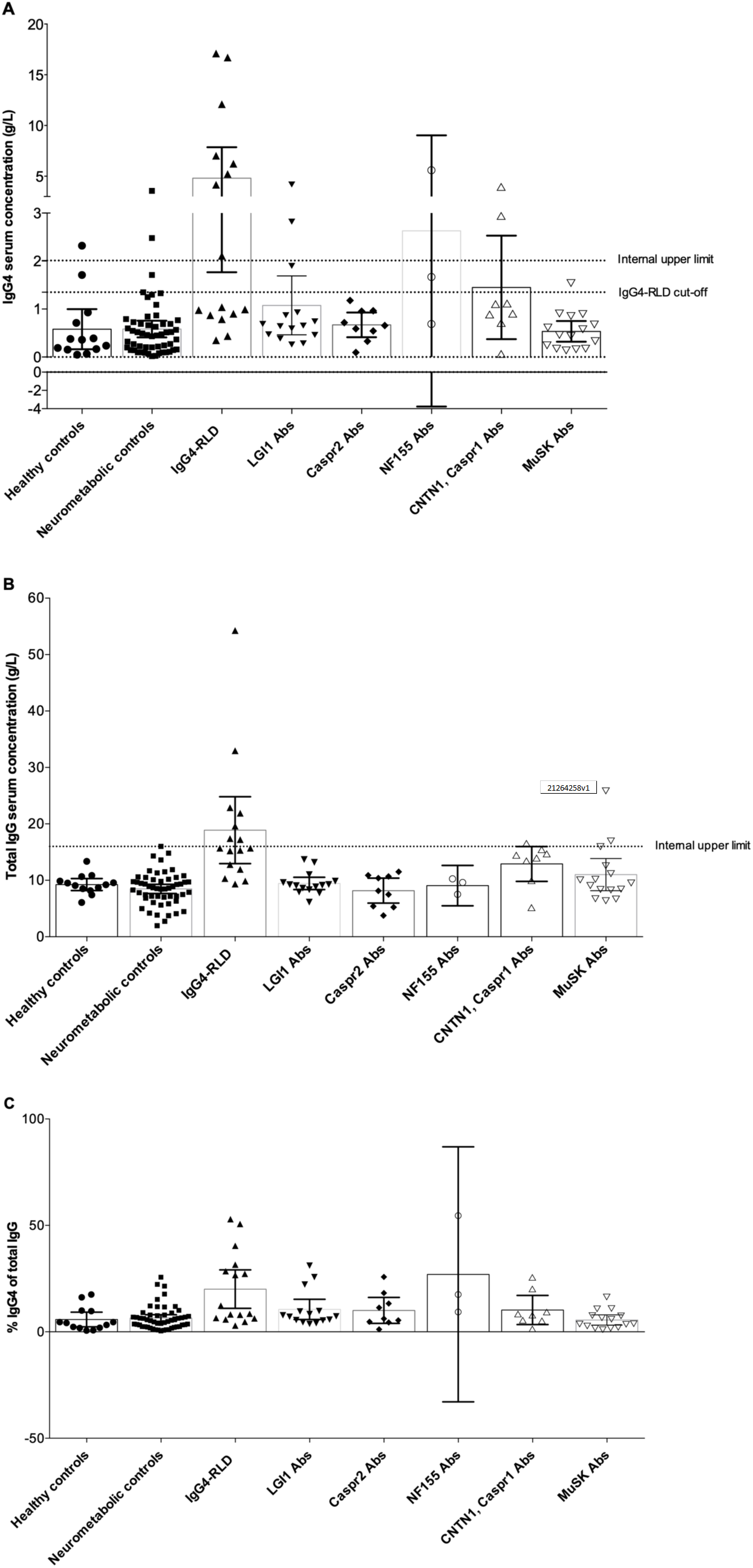
Serum IgG4 and total IgG concentrations in patients with IgG4 autoantibodies, IgG4-RLD, neurometabolic and healthy controls. IgG4 concentrations were obtained by nephelometry. **A)** Serum IgG4 concentrations. The internal upper limit for IgG4 concentrations is indicated as a line at 2.01 g/L, the official cut-off for elevated IgG4 concentrations in IgG4-RLD is indicated as a line at 1.35 g/L. **B)** Total serum IgG concentrations. The internal upper limit for total IgG is indicated as a line at 16 g/L. **C)** Percent IgG4 of total IgG. Bar graphs indicate mean and error bars indicate 95% CI. Abs = antibodies; Caspr1 = contactin-associated protein-like 1; Caspr2 = contactin-associated protein-like 2; CNTN1 = contactin 1; IgG = immunoglobulin type G; IgG4 = immunoglobulin type G subclass 4; IgG4-RLD = IgG4-related disease; LGI1 = leucine-rich glioma inactivated protein-1; MuSK = muscle-specific kinase; NF155 = neurofascin 155.

Patients with IgG4-RLD also had elevated total serum IgG concentrations (Figure 2B) and increased relative IgG4 concentrations (Figure 2C). We reasoned that in these patients, serum IgG4 contributed substantially to the total IgG concentrations, and we found a significant correlation between total IgG and IgG4 (r=0.9265, *p*<.0001, Figure 3C). Significant correlations were also observed in the neurometabolic controls (Figure 3B) and in patients with LGI1 autoantibodies (Figure 3D) with elevated IgG4 concentrations, but their total serum IgG concentrations were in the normal range (Figure 2B). No correlations were found in the other groups (Figure 3A, E, G, H).

**Figure 3:**
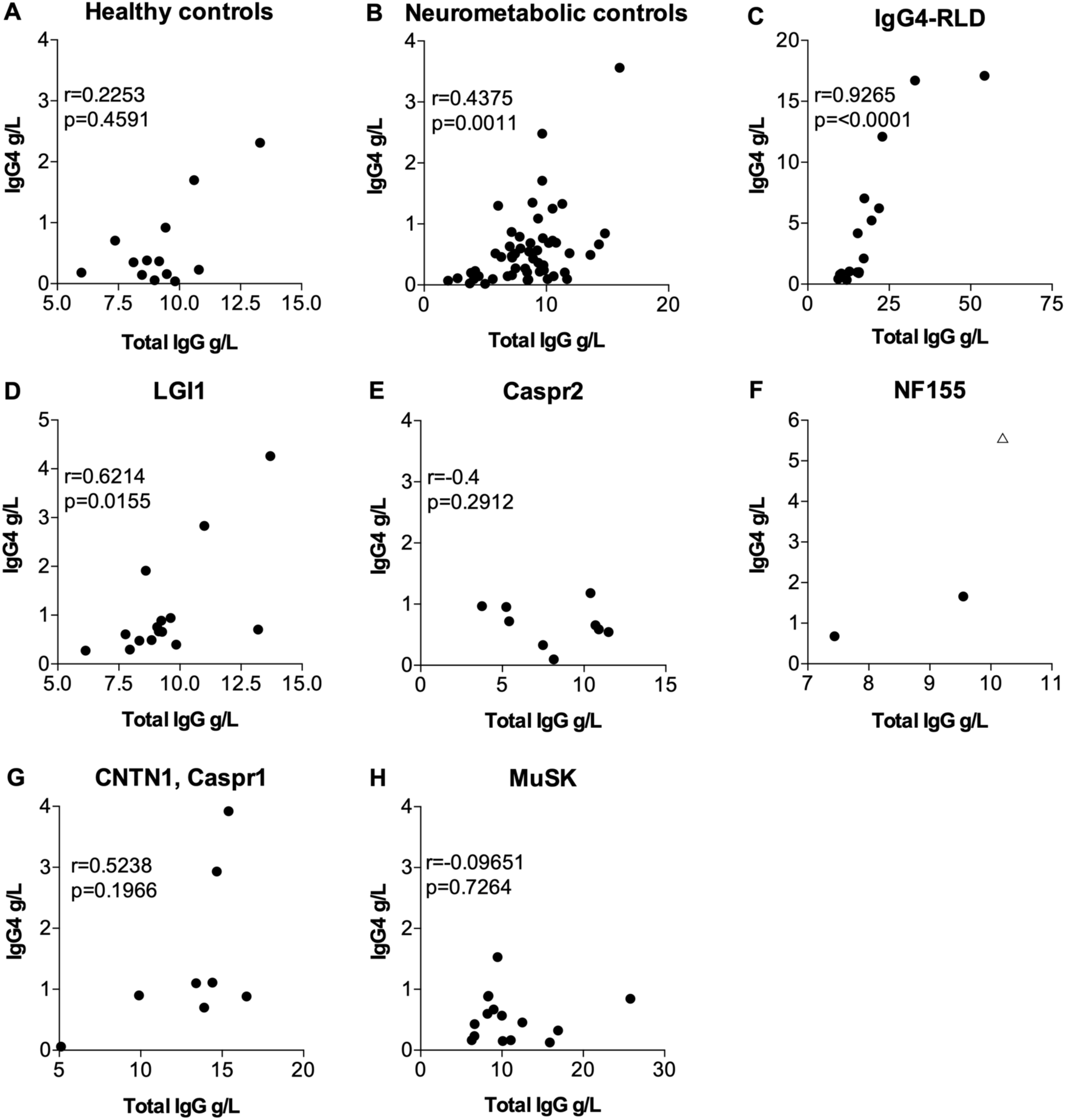
Correlation between serum IgG4 and total IgG concentrations (statistical analysis with Spearman correlation in datasets with at least 3 datapoints). **A)** healthy controls, **B)** neurometabolic controls, **C)** IgG4-RLD, **D)** LGI1, **E)** Caspr2, **F)** NF155, **G)** CNTN1, Caspr1 and **H)** MuSK. In **F)**, the triangular data point indicates a patient that was excluded from the statistical analysis due to the presence of pan-NF antibodies instead of NF155 antibodies and severe concomitant autoimmune diseases. Caspr1 = contactin-associated protein-like 1; Caspr2 = contactin-associated protein-like 2; CNTN1 = contactin 1; IgG = immunoglobulin type G; IgG4 = immunoglobulin type G subclass 4; IgG4-RLD = IgG4-related disease; LGI1 = leucine-rich glioma inactivated protein-1; MuSK = muscle-specific kinase; NF155 = neurofascin 155.

### Serum IgG4 concentrations were higher in males than in females

We observed that IgG4 was more frequently elevated in males than in females (Table 1). In IgG4-RLD, IgG4 was elevated in 85.71% of males but only in 22.22% of females. These different proportions were significant in a Fisher’s exact test (likelihood ratio: 6.904, *p* = .041). Elevated IgG4 was also observed mainly in male patients with IgG4-AID and their absolute IgG4 concentrations were higher than in females (Figure 4), with maximum values of 17.1 g/L male vs. 12.1 g/L female (IgG4-RLD), 5.55 g/L male vs. 1.53 g/L female (IgG4-AID), 3.56 g/L vs. 1.71 g/L female (neurometabolic controls) and 2.31 g/L male vs. 0.92 g/L female (healthy controls).

**Figure 4:**
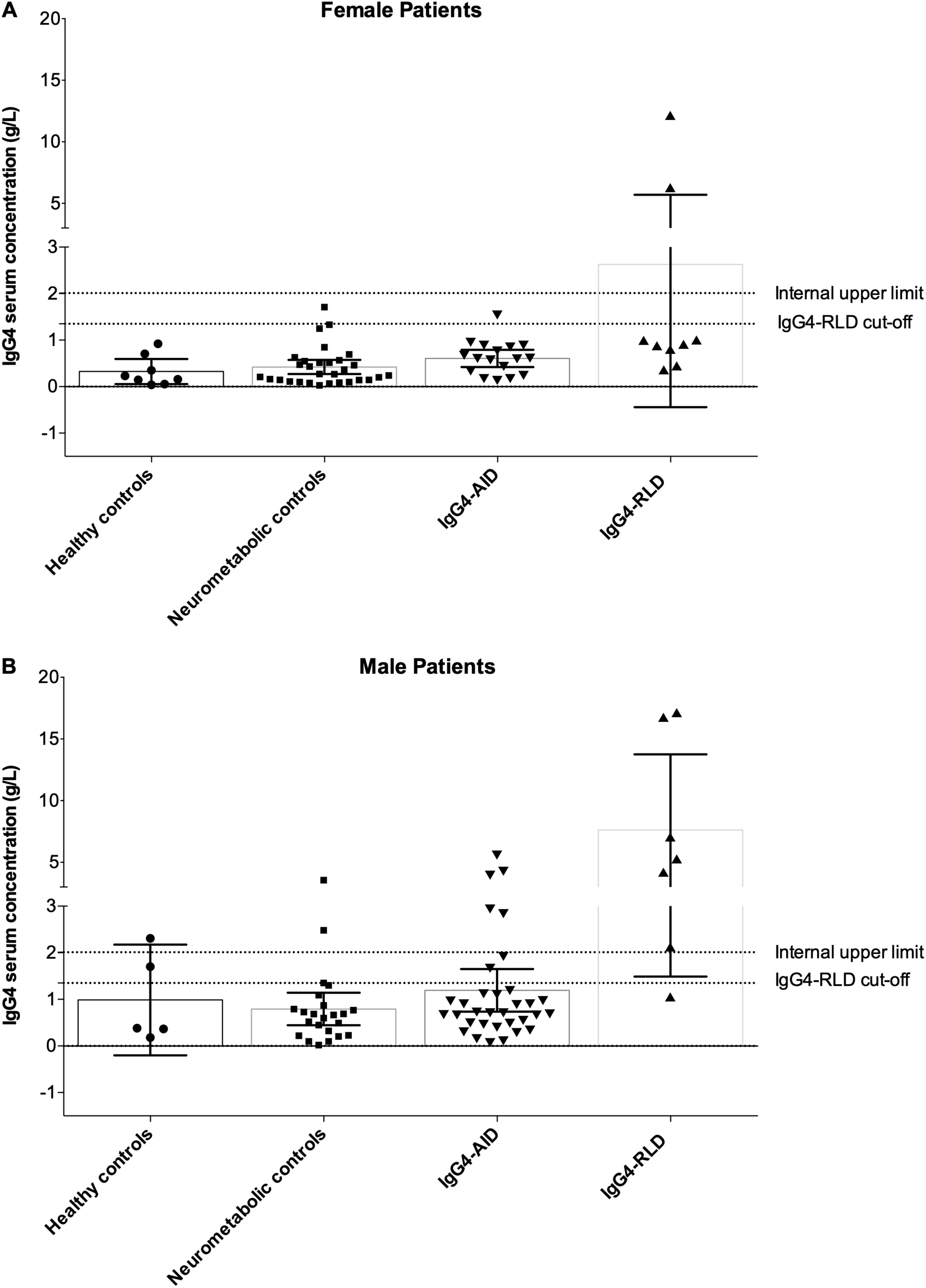
Gender-specific serum IgG4 concentrations. **A)** Serum IgG4 concentrations in female patients, **B)** Serum IgG4 concentrations in male patients. Bar graphs indicate mean, error bars indicate 95% CI. IgG4 = immunoglobulin type G subclass 4; IgG4-AID = IgG4 autoimmune disease; IgG4-RLD = IgG4-related disease.

### Serum IgG4 levels did not correlate with anti-neuronal/neuromuscular autoantibody titres

We further addressed whether serum IgG4 concentrations were associated with anti-neuronal/neuromuscular autoantibody titres, but found no overall correlation (Supplementary Figure 1). Interestingly, one patient with pan-NF antibodies showed a highly elevated relative and absolute serum IgG4 concentration (5.55 g/L, 54% IgG4 of total IgG) and an exceptionally high serum antibody titre of 1:40,960. Nevertheless, this patient had severe comorbidities for 11 years including multiple sclerosis and Grave’s disease, and was treated with intravenous immunoglobulin (IVIg), plasma exchange (PLEX) and Interferon β1a and was therefore excluded as outlier from the statistics.

### Lack of overlap between IgG4-AID and IgG4-RLD

16% of the IgG4-AID patients showed increased IgG4 serum concentrations (Table 1). Their clinical and histopathological data were analysed for key symptoms of IgG4-RLD that are considered as diagnostic criteria for IgG4-RLD (2, 4), specifically 1) organ enlargement, 2) tumefactive lesions, 3) fibrosis or 4) IgG4^+^ plasma cell infiltrates (Figure 5A-D). None of the IgG4-AID patients, including those with elevated IgG4 serum levels, fulfilled these diagnostic criteria for IgG4-RLD.

**Figure 5:**
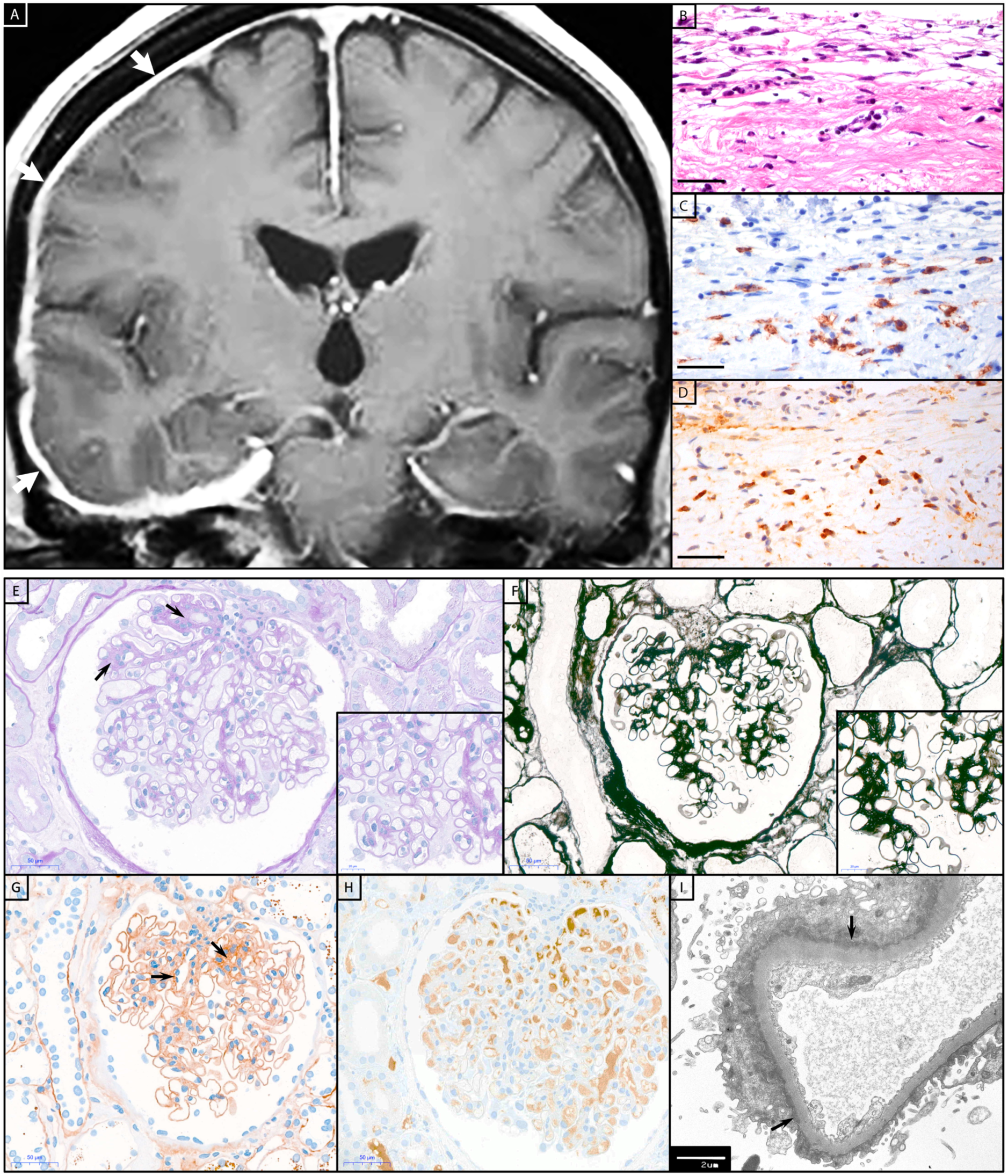
Histopathology of IgG4-RLD and IgG4-AID patients. Patient with IgG4-RLD: **A)** MRI shows a right-sided thickening and increased contrast agent uptake of the pachymeninges (white arrows). A biopsy from the meninges reveals fragments of dura with **B)** prominent fibrosis and infiltration with **C)** numerous CD138^+^ and **D)** IgG4^+^ plasma cells, compatible with IgG4-RLD pachymeningitis. Patient with IgG4-AID: A kidney biopsy from a patient with CNTN1/Caspr1-complex autoantibodies and acute kidney failure, nephrotic syndrome, hypoalbuminemia and microhaematuria. **E)** Glomerulus with mild segmental fibrous mesangial expansion (arrows), somewhat thickened capillary basal membranes, without hypercellularity (PAS stain). **F)** Silver stain with smooth capillary loop basal membranes. **G)** Immunohistochemistry for IgG with global finely granular peripheral and segmental mesangial positive deposits (arrows). **H)** Negative immunohistochemistry for IgG4. **I)** Electron microscopy with numerous small sub-epithelial electron-dense deposits with flattening of podocytic foot processes (arrows). Scale bar B-D, E-H = 50 µm; scale bar inset in E and F = 20 µm; scale bar I = 2 µm.

Neuronal proteins such as CNTN1 and neurofascin186 are also expressed on podocytes in the kidney (Figure 6, (10, 11)). Accordingly, patients with chronic inflammatory demyelinating polyradiculoneuropathy (CIDP) may develop membranous glomerulonephritis (12-20). Therefore, the kidney is also a target for IgG4 autoantibodies in these patients (21, 22). Histopathological characteristics of IgG4-RLD were investigated in a kidney biopsy of one CIDP patient with CNTN1/Caspr1-complex autoantibodies, nephrotic syndrome and microhaematuria.

**Figure 6:**
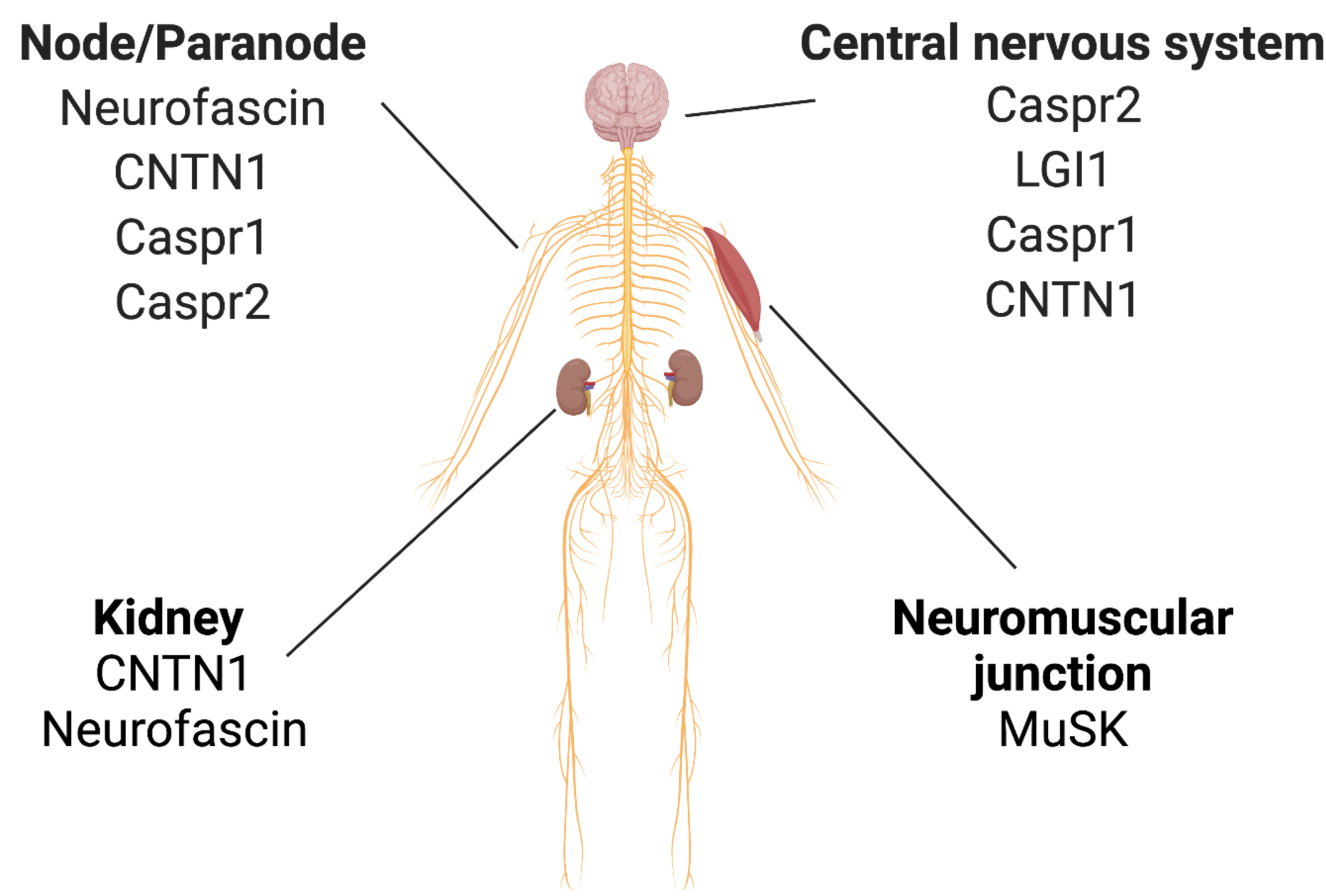
Expression of selected neuronal/neuromuscular antigens that are targeted by pathogenic IgG4 autoantibodies (in a selection of relevant organs). Caspr1 = contactin-associated protein-like 1; Caspr2 = contactin-associated protein-like 2; CNTN1 = contactin 1; LGI1 = leucine-rich glioma inactivated protein-1; MuSK = muscle-specific kinase.

The histopathological workup of the kidney biopsy (Figure 5E-I) showed minimal membranous glomerulopathy class I with normal glomeruli by light microscopy but mesangial immune deposits by electron microscopy. Nevertheless, neither IgG4 deposits, fibrosis nor IgG4^+^ plasma cell infiltrates were observed in this biopsy.

We further addressed whether IgG4-RLD patients display anti-neuronal/neuromuscular autoantibodies. Sera from nine patients with IgG4-RLD were available and were tested in tissue-based assays on rat brain for the presence of anti-neuronal/neuromuscular autoantibodies. All sera were negative in the tissue-based assay (data not shown).

## Discussion

IgG4-AID and IgG4-RLD are two groups of rare diseases associated with IgG4. To date, it has not been systematically analysed whether these two groups are part of the same disease spectrum. To the best of our knowledge, we are the first to answer this question by performing a comparative analysis between neuronal/neuromuscular cell surface IgG4 autoimmunity and IgG4-RLD to see if they share diagnostic characteristics.

As a result, we could not observe any indication for an overlap between anti-neuronal IgG4-AID and IgG4-RLD, as firstly, 84% of patients with IgG4-AID had normal IgG4 levels, and serum IgG4 concentrations did not correlate with antigen specific autoantibody titres and, secondly, we could not identify clinical or histopathological indications for IgG4-RLD, while, thirdly, a substantial fraction of IgG4-RLD patients (50%) had elevated IgG4 serum concentrations, but all IgG4-RLD patients were negative for anti-neuronal/neuromuscular autoantibodies. In a tissue-based assay, which represents a broad screening method for autoantibodies against a variety of neuronal and glial epitopes, no reactivity was found in the sera from patients with IgG4-RLD. However, we did not test for the presence of non-neuronal IgG4 autoantibodies, which therefore cannot be ruled out.

In the healthy and neurometabolic controls as well as in the IgG4-AID patient group, few individuals showed elevated serum IgG4 concentrations. This has to be expected, since IgG4 concentrations vary and may temporarily/seasonally increase due to a change in immune status, e.g. due to infections or allergy. IgG4 levels are known to be higher in males than in females (23, 24), which we also observed, with most male IgG4-RLD patients (85.71%) presenting with elevated serum IgG4 in contrast to only 22.22% of female IgG4-RLD patients. This predisposition to elevated serum IgG4 may pose male patients at increased risk for suffering from IgG4-RLD, which accord to the male predominance of IgG4-RLD (Table 2). Elevated IgG4 levels may offer better protection from classical IgG1/ IgG3 mediated autoimmune diseases such as AChR myasthenia gravis (MG), which is indeed more frequent in women (25). Interestingly, AChR-IgG4 protected from MG in an animal model (26). Nevertheless, pathogenic IgG4 autoantibodies cause pemphigus and MuSK myasthenia gravis, which have a clear female predominance (Table 2). However, we found that the majority of MuSK MG patients had normal/low serum IgG4 concentrations, suggesting that the total IgG4 concentrations are unrelated to MuSK antibody pathogenicity. Our observations were in line with another study that describes normal IgG4 levels in the majority of pemphigus patients (24/27 pemphigus vulgaris and 13/16 pemphigus foliaceus patients had normal IgG4 levels) (27).

**Table 2:** Summary of IgG4-AID vs. IgG4-RLD. † Few epidemiological data available, the values shall be considered as estimates. ‡ characteristics are considered as pathognomonic. Caspr2 = contactin-associated protein-like 2; CIDP = chronic inflammatory demyelinating polyneuropathy; CNTN1 = contactin 1; HLA = human leucocyte antigen; IgG4 = immunoglobulin type G subclass 4; LGI1 = leucine-rich glioma inactivated protein-1; MG = myasthenia gravis; MuSK = muscle-specific kinase; NF155 = neurofascin 155.

| Disease aspects | IgG4 autoimmune diseases | IgG4-related diseases | References |
| --- | --- | --- | --- |
| Prevalence†, (per 10,000) | <0.0001 – 5 | <0.01 - 8 | orpha.net |
| Gender predisposition†, | MuSK MG, pemphigus, thrombotic thrombocytopenic purpura: female predominance<br>LGI1, Caspr2 encephalitis, CIDP (NF155, CNTN1): male predominance | Male predominance | [20-24] |
| Affected organs | Currently known: nervous system, kidneys, blood, skin and mucosa | All organs / multiorgan, often in salivary glands, lymph nodes and pancreas | [3, 25] |
| Fibrosis | No | Yes ‡ | [25] |
| Tissue infiltrates of IgG4<sup>+</sup> lymphocytes | No | Yes ‡ | [25] |
| Organ enlargement, tumour-like mass formation in affected organ | No | Yes, often in lacrimal glands, orbits, major salivary glands, pancreas, bile ducts, retroperitoneum, lungs, kidneys, aorta, pachymeninges and thyroid gland | [25] |
| Suspected HLA risk loci | HLA-DRB1*14, HLA-DQB1*05, HLA-DRB1*14-DQB1*05, HLA-DRB1*15, HLA-DRB1*04,<br>DRB1*03 protective | HLA-DRB1*04:05, HLA-DQB1*04:01, HLA-A, HLA-C, HLA I, HLA-DQB1*03:02, HLA-B*07, HLA-B*08, HLA-DRB1*15 | [26, 27] |
| IgG4 concentrations | Normal | Elevated ($\geq 1.35$g/L in 70% of patients) ‡ | [28] |
| Autoantigen-specific IgG4 | In 100% of cases ‡ | In a subset of patients | [3, 29] |
| Location of known IgG4 autoantigen | Extracellular | Intracellular and extracellular | [3, 29] |
| Role of IgG4 | Directly pathogenic ‡ | Unclear | [3, 29] |
| Pathogenic mechanism of IgG4 | IgG4 blocks protein-protein interactions | Unknown | [3, 29] |
| Treatment response | Moderate success of corticosteroid treatment. B cell depletion beneficial especially in treatment resistant patients | Moderate success of corticosteroid treatment. B cell depletion beneficial especially in treatment resistant patients | [30, 31] |

Importantly, antigen-specific IgG4 directly cause neurological symptoms of IgG4-AID (28), while the pathogenic mechanisms of IgG4 in IgG4-RLD are currently not well understood. So far, only very few target antigens have been described in IgG4-RLD (29), but these are mostly located intracellularly. For example, antibodies in IgG4-related autoimmune pancreatitis (IgG4-AIP), a form of IgG4-RLD, may target annexin A11 (30), which is located in the nucleus (31). Passive transfer of patient IgG1 and IgG4 from patients with IgG4-AIP to experimental animals^2^ showed that both IgG1 and IgG4 induced pancreatic injury, but IgG4 also led to significant reduction of necrosis when co-injected with IgG1 (32). Similar observations were made with IgG4 against annexin A11, which blocked binding of IgG1 (30). Therefore, at the moment the role of IgG4 in IgG4-RLD remains elusive.

Interestingly, some overlap of IgG4-AID and IgG4-RLD is currently discussed for anti-neutrophil cytoplasmic autoantibodies (ANCA) associated vasculitis (granulomatosis with polyangiitis; GPA; also called Wegener’s granulomatosis). GPA is characterized by antigen-specific IgG4, IgG3 or IgG1 against proteinase 3 (PR3; surface antigen) or myeloperoxidase (MPO; intracellular protein) in neutrophils and monocytes, increased levels of IgG4^+^ plasma cells, fibrosis and sometimes elevated serum IgG4 levels (33-36). The patients may additionally present with IgG4-RLD (37-39), but also with PLA2R autoantibodies (40), which are found in another IgG4-AID, PLA2R-antibody positive membranous nephropathy (41). Nevertheless, it is still unclear whether GPA belongs to the IgG4-AID, as the pathogenicity of IgG4-PR3 has not yet been demonstrated by passive transfer of IgG4 to experimental animals.

## Conclusion

IgG4-AID and IgG4-RLD are most likely distinct disease groups. Due to their low disease prevalences, comparative data to characterize these diseases are limited. In our study we provide three relevant findings, 1) a significantly higher proportion of IgG4-RLD patients (50%) had elevated serum IgG4 concentrations compared to IgG4-AID (16%), 2) IgG4-AID patients with elevated IgG4 did not meet the diagnostic criteria of IgG4-RLD and their autoantibody titres did not correlate with serum IgG4 concentrations, while 3) patients with IgG4-RLD were negative for anti-neuronal/neuromuscular IgG4 autoantibodies. Furthermore, male IgG4-RLD patients presented significantly more frequently with elevated serum IgG4 compared to female patients.

In summary, our data do not support clinical or histopathological commonalities between IgG4-AID and IgG4-RLD, suggesting that they are in fact unrelated. Further studies on IgG4-AID and IgG4-RLD will lead to a better understanding of these diseases.

## Data Availability

The data that support the findings of this study are available from the corresponding author upon reasonable request.

## Acknowledgements

We thank our colleagues Irene Erber and Anita Krnjic for excellent technical assistance.

## Funding

This work was supported by grants from the Austrian Science Fund (FWF), project number T996-B30, SYNABS project number I4685-B, DOC33-B27 and the Austrian Society of Neurology (Österreichische Gesellschaft für Neurologie). Figure 6 was created with BioRender software (license IK).

## Conflicts of interest

V. Endmayr reports no disclosures relevant to the manuscript.

C. Tunc reports no disclosures relevant to the manuscript.

L. Ergin reports no disclosures relevant to the manuscript.

A. de Rosa reports no disclosures relevant to the manuscript.

R. Weng reports no disclosures relevant to the manuscript.

L. Wagner reports no disclosures relevant to the manuscript.

T.Y. Yu reports no disclosures relevant to the manuscript.

A. Fichtenbaum reports no disclosures relevant to the manuscript.

T. Perkmann reports no disclosures relevant to the manuscript.

H. Haslacher reports no disclosures relevant to the manuscript.

N. Kozakowski reports no disclosures relevant to the manuscript.

C. Schwaiger reports no disclosures relevant to the manuscript.

G. Ricken reports no disclosures relevant to the manuscript.

S. Hametner reports no disclosures relevant to the manuscript.

L. Almeida Dutra received a grant from Fleury Laboratory for the Brazilian Autoimmune Encephalitis Project without personal compensation.

C. Lechner served as a consultant for Roche.

D. de Simoni reports no disclosures relevant to the manuscript.

K.N. Poppert received a travel grant from Merck.

G.J. Müller reports no disclosures relevant to the manuscript.

S. Pirker reports no disclosures relevant to the manuscript.

W. Pirker reports no disclosures relevant to the manuscript.

A. Angelovski reports no disclosures relevant to the manuscript.

M. Valach reports no disclosures relevant to the manuscript.

M. Maestri reports no disclosures relevant to the manuscript.

M. Guida reports no disclosures relevant to the manuscript.

R. Ricciardi reports no disclosures relevant to the manuscript.

F. Frommlet reports no disclosures relevant to the manuscript.

D. Sieghart reports no disclosures relevant to the manuscript.

M. Pinter reports no disclosures relevant to the manuscript.

R. Höftberger reports speakers’ honoraria from Novartis and Biogen. The Medical University of Vienna (Austria; employer of Dr. Höftberger) receives payment for antibody assays and for antibody validation experiments organized by Euroimmun (Lübeck, Germany).

I. Koneczny reports no disclosures relevant to the manuscript.

## Footnote

1 www.orpha.net

2 If the pathogenic mechanism depends on interaction with other parts of the immune system, precautions such as injection of human complement along with the antibodies are necessary as human and animal immune systems are not always compatible.

