## Supplementary material for "Anti-neuronal IgG4 autoimmune diseases and IgG4-related diseases may not be part of the same spectrum: a comparative study"

### Supplementary Figure 1

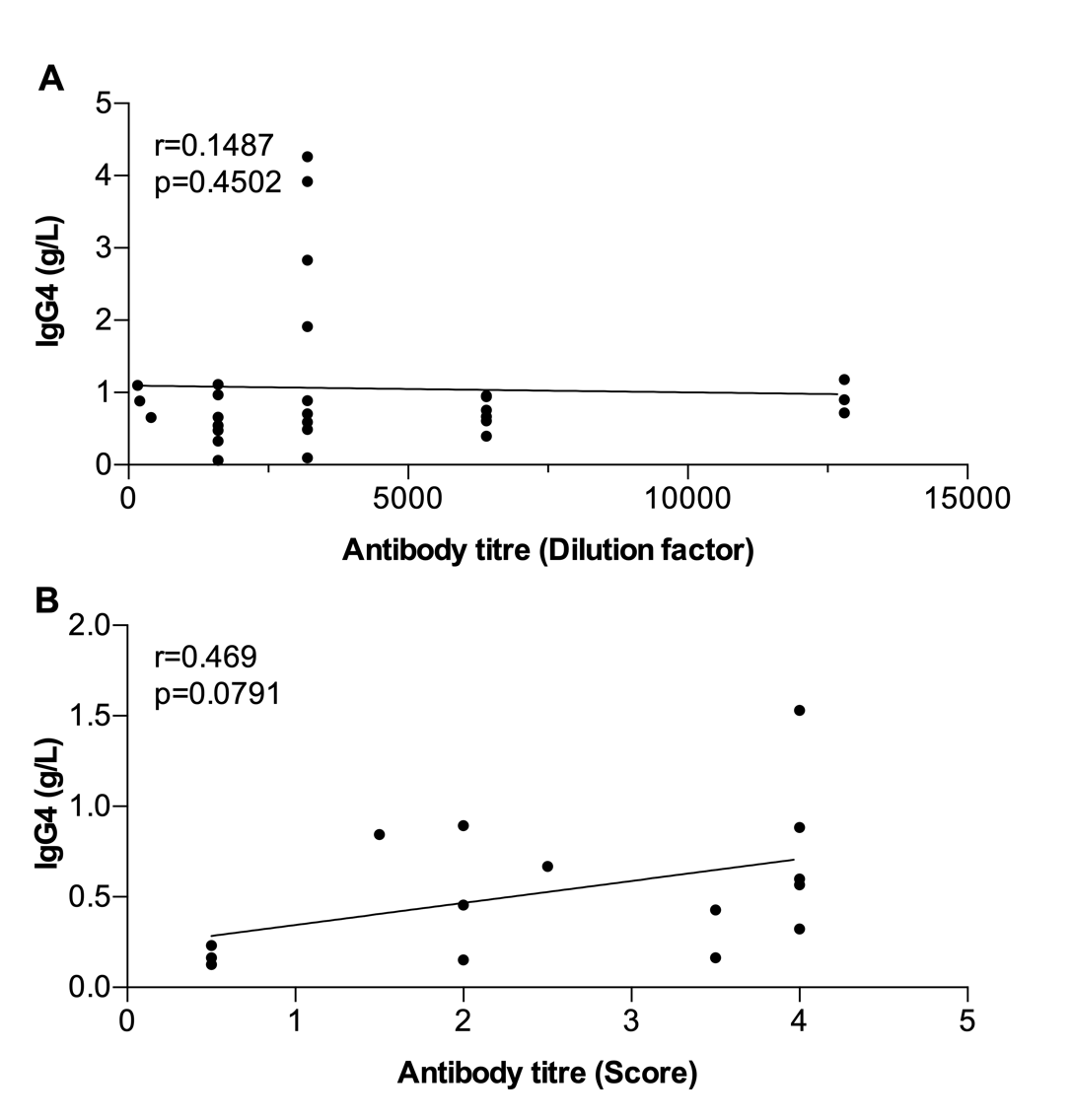

Supplementary Figure 1: No apparent correlation between serum IgG4 concentrations and IgG4 autoantibody titres could be observed. (A) LGI1, Caspr2, NF155, CNTN1/Caspr1 antibody titres are expressed as dilution factor at which autoantibodies were still detectable. (B) MuSK antibody titres are determined by visual scoring of immunofluorescent signal in cell-based assay, from 0 = negative to 4 = highest intensity. Nonparametric Spearman correlation with two-tailed p-value. IgG4 = immunoglobulin type G subclass 4.

**Supplementary Table 1: Clinical and epidemiological data of the study cohort.**

AChR = acetylcholine receptor; AIE = autoimmune encephalitis; approx. = approximately; Aza = azathioprine; Caspr1 = contactin-associated protein-like 1; Caspr2 = contactin-associated protein-like 2; CIDP = chronic inflammatory demyelinating polyneuropathy; CNTN1 = contactin 1; CSF = cerebrospinal fluid; d = day; F = female; GBS = Guillain-Barré syndrome; HDMP = high dose methylprednisolone; IG = immunoglobulin; IgG = immunoglobulin type G; IgG4 = immunoglobulin type G subclass 4; IgG4-RLD = immunoglobulin G4-related disease; IVIg = intravenous immunoglobulins; i.v. = intravenous; LE = lower extremities; LGI1 = leucine-rich glioma inactivated protein- 1; M = male; MG = myasthenia gravis; min = minutes; M. levator palpebrae sup. = musculus levator palpebrae superioris; MRI = magnetic resonance imaging; mRS = modified rankin scale; M. rectus sup. = musculus rectus superior; MS = multiple sclerosis; MuSK = muscle-specific kinase; n.a. = not applicable; neg. = negative; NF155 = neurofascin155; pan-NF = positive for pan neurofascin antibodies (155/140/186); PLEX = plasma exchange; Prs = prednisolone; p.o. = per os; Pyr = pyridostigmine bromide; RTX = rituximab; sc = subcutaneous; UE = upper extremities.

| **patient ID** | **antigen** | **sex** | **clinical diagnosis** | **clinical symptoms** | **CSF [cell/µL]** | **oligoclonal bands** | **highest mRS** | **acute therapy** |
| --- | --- | --- | --- | --- | --- | --- | --- | --- |
| LGI1 #1 | LGI1 | F | AIE | MRI-neg. focal epilepsy: several episodes of blurry vision, general weakness, impaired mentation of approx. 10-15 min duration per day, approx. 7 episodes of electrifying paraesthesias and generalized goose bumps of 3-4 seconds duration per day | n.a. | n.a. | 1 | lamotrigine 100 mg/d |
| LGI1 #2 | LGI1 | M | AIE | anxiety, cognitive impairment, autonomic disturbance, stereotypical behaviour | 3 | neg. | 2 | HDMP, IVIg, RTX, Prs p.o. |
| LGI1 #3 | LGI1 | M | AIE | fasciculations of the lower leg, dysaesthesia of the lateral thigh area, insomnia, memory impairment | 2 | n.a. | 1 | HDMP, IVIg |
| LGI1 #4 | LGI1 | F | AIE | anxiety, seizures, cognitive impairment, nausea | 2 | neg. | 1 | IVIg, RTX |
| LGI1 #5 | LGI1 | M | AIE | psychosis, anxiety, insomnia, social withdrawal, seizures, cognitive impairment, dyskinesia, autonomic disturbance, recurrent diarrhoea for few weeks to months | 2 | neg. | 2 | HDMP, IVIg, RTX, Prs p.o. |
| LGI1 #6 | LGI1 | M | AIE | psychosis | 7 | neg. | 1 | HDMP, RTX, Prs p.o. |
| LGI1 #7 | LGI1 | M | AIE | reduced vigilance, cognitive impairment, autonomic disturbance | 0 | n.a. | 1 | IVIg, Prs p.o. |
| LGI1 #8 | LGI1 | M | AIE | psychosis, anxiety, insomnia, social withdrawal, cognitive impairment | 3 | n.a. | 2 | HDMP, Prs p.o. |
| LGI1 #9 | LGI1 | M | AIE | cognitive impairment | 1 | neg. | 1 | Prs p.o. |
| LGI1 #10 | LGI1 | F | AIE | MRI-neg. focal epilepsy, cognitive impairment, depression, hallucinations, frequent subclinical temporal lobe seizures, pressing occipital headaches of location, lightheadedness, general weakness, impaired mentation, frequent trance-like episodes with formed visual hallucinations, jerking of right UE, staring gaze, oral automatisms and altered breathing pattern of few seconds duration | 0 | neg. | 3 | HDMP, 2 cycles of IVIg, 4 cycles of RTX |
| LGI1 #11 | LGI1 | M | AIE | social withdrawal, autonomic disturbance, crampi, fasciculations, myokymia | 13 | neg. | 1 | IVIg, Prs p.o. |
| LGI1 #12 | LGI1 | F | AIE | memory disturbances, confusion, epilepsy, inattention, hyponatremia of 118 mmol/L | 0.3 | n.a. | n.a | valproate, quetiapine, HDMP, IVIg |
| LGI1 #13 | LGI1 | M | AIE | focal seizures, insomnia, memory impairment, pallhypaesthesia, hyporeflexia, cognitive impairment, personality change | 26 | neg. | 0 | HDMP, RTX, Prs p.o. |
| LGI1 #14 | LGI1 | M | AIE | agitation, insomnia, memory and language disturbances, confusion, epilepsy, hyponatremia of 116 mmol/L | 1 | n.a. | n.a. | escitalopram 10 mg/d, sodium replacement, levetiracetam 1000 mg/d, lacosamide 200 mg/d, HDMP |
| LGI1 #15 | LGI1 | M | AIE | focal seizures, vertigo symptoms, epigastic auras | 1 | neg. | 0 | HDMP, IVIg |
| Caspr2 #1 | Caspr2 | M | AIE | seizures, speech disturbance | 3 | neg. | 1 | IVIg, RTX, Prs p.o. |
| Caspr2 #2 | Caspr2 | M | small fibre neuropathy | neuropathic pain | n.a. | n.a. | n.a | - |
| Caspr2 #3 | Caspr2 | M | AIE | limbic encephalitis | n.a. | n.a. | n.a. | - |
| Caspr2 #4 | Caspr2 | M | AIE | reduced vigilance, cognitive impairment, dyskinesia | 12 | n.a. | 3 | HDMP, Prs p.o. |
| Caspr2 #5 | Caspr2 | M | AIE | psychosis, memory deficits, sensory ataxia with neuropathic pain | 4 | neg. | 2 | HDMP, Prs p.o. |
| Caspr2 #6 | Caspr2 | M | AIE | cognitive impairment, organic psychosyndrome | 0 | neg. | 0 | HDMP, RTX, PLEX, Prs p.o. |
| Caspr2 #7 | Caspr2 + LGI1 + AchR | M | AIE | behavioural, psychiatric and cognitive disturbances, movement disorder, tremor, severe episodes of pain on the extremities | 1 | neg. | n.a. | thymectomy |
| Caspr2 #8 | Caspr2 | M | AIE | agitation, confusion, dyskinesia, abnormal posture, orofacial dyskinesia, autonomic instability, sleep impairment, heteroaggressivity, auditory hallucinations, mood alteration, carrier of membranous nephropathy since 2016 | 0.3 | n.a. | n.a. | no immunosuppressive therapy |
| Caspr2 #9 | Caspr2 | F | AIE | rare myokymias, fasciculations, cramps | n.a. | n.a. | n.a. | thymoma (8 years ago) treated with surgery, chemotherapy and radiotherapy / no immunosuppressive therapy |
| NF155 #1 | NF155 | M | CIDP with minimal symptoms | sensory motor neuropathy of UE and LE, tremor | 10 | yes | 1 | IVIg |
| NF155 #2 | NF155 | M | CIDP | ataxia, tremor, sensory deficits of UE and LE | 21 | neg. | 5 | IVIg, RTX (two 1g doses 2 weeks apart), PLEX, Prs p.o. |
| NF155 #3 | pan-NF155/140/186 | M | GBS + MS (since 2010) + Graves´ disease | tetraparesis, sensory motor deficits of UE and LE; death | 1 | n.a. | 5 | IVIg, RTX (two 1g doses 2 weeks apart), PLEX |
| CNTN1 #1 | CNTN1 + Caspr1 | M | CIDP | oculomotor nerve paresis, ptosis, ataxia, diplopia, dysphagia, tetraparesis, intubation; death | n.a. | n.a. | n.a. | IVIg, PLEX |
| CNTN1 #2 | CNTN1 + Caspr1 | M | CIDP | senso-motoric neurological deficits of UE and LE, sensory ataxia, dysesthesia, edema (ascites and leg edema), hypoproteinemia, hypoalbuminemia, proteinuria | normal (<5) | neg. | n.a. | i.v. methylprednisolone (4x 250 mg and 3x 125 mg, over 7 d), followed by Prs p.o. |
| CNTN1 #3 | CNTN1 | M | CIDP | - | n.a. | n.a. | n.a. | 3 cycles of IVIg |
| CNTN1 #4 | CNTN1 | M | CIDP | - | n.a. | n.a. | n.a. | IVIg |
| CNTN1 #5 | CNTN1 | M | acute onset CIDP | - | n.a. | n.a. | n.a. | - |
| CNTN1 #6 | CNTN1 | M | CIDP | gait disorder, paresthesias in stocking-glove distribution, intention tremor, weak tendon reflexes UE, absent tendon reflexes LE | 5 | neg. | 4 | 2 cycles of IVIg |
| CNTN1 #7 | CNTN1 | M | CIDP | ascending paraparesis, diplopia, facial paralysis right | 4 | n.a. | 0 | IVIg, RTX, PLEX |
| CNTN1 #8 | Caspr1 | M | CIDP | tetraparesis, stocking-glove distribution hypesthesia, absent tendon reflexes | 6 | neg. | 5 | 3 cycles of IVIg, PLEX (1 cycle of 5 sessions), RTX |
| MuSK #1 | MuSK | F | generalised MG (initially only ocular symptoms) | - | - | - | 0 | thymectomy / Pyr, Prs, Aza |
| MuSK #2 | MuSK | F | generalised MG (initially only ocular symptoms) | ophtalmoparesis, generalized muscle weakness | - | - | 1 | Pyr, Prs, Aza, RTX |
| MuSK #3 | MuSK | F | generalised MG | dysarthria, dysphagia, ophtalmoparesis, ptosis, facial muscle weakness | - | - | 2 | RTX |
| MuSK #4 | MuSK | M | generalised MG | - | - | - | 0 | Pyr, Prs, Aza |
| MuSK #5 | MuSK, borderline | M | ocular MG | - | - | - | 1 | Prs p.o. |
| MuSK #6 | MuSK | M | generalised MG | ptosis, ophtalmoparesis, proximal muscle weakness, dysarthria, dysphagia | - | - | n.a. | Pyr, Prs, IVIg, RTX, Ibrutinib |
| MuSK #7 | MuSK | F | generalised MG | orofacial muscle weakness, dysarthria | - | - | 1 | - |
| MuSK #8 | MuSK | F | generalised MG | dysarthria, dysphagia, ophtalmoparesis, ptosis, orofacial and extremity muscle weakness, dyspnea | - | - | 2 | thymectomy / Pyr, Prs, sc IG, RTX |
| MuSK #9 | MuSK | F | generalised MG | ophtalmoparesis, dysphagia, proximal muscle weakness | - | - | 1 | Pyr, RTX |
| MuSK #10 | MuSK | F | generalised MG | dyspnea, dysphagia, dysarthria, proximal muscle weakness | - | - | 2 | thymectomy / Pyr, Prs, Aza, RTX |
| MuSK #11 | MuSK | F | generalised MG | - | - | - | 2 | thymectomy / Prs, IVIg |
| MuSK #12 | MuSK | F | generalised MG | dysarthria, dysphagia, ophtalmoparesis | - | - | 2 | Prs, RTX |
| MuSK #13 | MuSK | F | generalised MG | - | - | - | 2 | thymectomy / Prs p.o. |
| MuSK #14 | MuSK | F | ocular MG | - | - | - | 1 | Pyr, Prs p.o. |
| MuSK #15 | MuSK | F | generalised MG | - | - | - | 1 | thymectomy/ IVIg, Prs p.o., cyclosporine |
| IgG4 RLD #1 | - | F | IgG4-related pachymeningitis | right frontal headache, diplopia with right-sided trochlear palsy, autoimmune diabetes, dura biopsy: fibrosis and IgG4^+^ plasma cells | 27 | yes | 2 | Prs p.o., 2 cycles of tocilizumab, 3 cycles of RTX |
| IgG4 RLD #2 | - | F | IgG4-RLD | elevated IgG4 levels in the pseudotumour | - | - | - | - |
| IgG4 RLD #3 | - | M | IgG4-RLD | chronic sclerosing sialadenitis (parotid gland) with up to 100 IgG4^+^ plasma cells/high power field | - | - | - | - |
| IgG4 RLD #4 | - | M | IgG4-RLD | bilateral swelling of the parotid | - | - | - | - |
| IgG4 RLD #5 | - | F | IgG4-RLD | pseudotumor orbitae, histology IgG4 associated disease | - | - | - | - |
| IgG4 RLD #6 | - | F | IgG4-RLD | swelling of glandula submandibularis, fibrosed adipose tissue with IgG4^+^ plasma cells increase and increased IgG4 to IgG ratio (70%) | - | - | - | - |
| IgG4 RLD #7 | - | F | IgG4-RLD | autoimmune pancreatitis, problems with salivary glands, biopsy of lacrimal gland compatible with IgG4-associated chronic dacryoadenitis | - | - | - | - |
| IgG4 RLD #8 | - | M | IgG4-RLD | chronic follicular sialadenitis with progressive glandular atrophy, partial sclerosis and significantly IgG4^+^ plasma cells | - | - | - | - |
| IgG4 RLD #9 | - | F | IgG4-RLD | axillary lymphadenopathy, swelling of the lacrimal glands | - | - | - | - |
| IgG4 RLD #10 | - | M | IgG4-RLD | orbita biopsy: pos. IgG4 staining | - | - | - | - |
| IgG4 RLD #11 | - | F | IgG4-RLD | muscle biopsy: M. levator palpebrae sup.: endomysial fibrosis, M. rectus sup.: T and B cells reactivity against IgG4 | - | - | - | - |
| IgG4 RLD #12 | - | M | IgG4-RLD | IgG4-RLD of the salivary gland | - | - | - | - |
| IgG4 RLD #13 | - | F | IgG4-RLD | thyroiditis, fibrosing variant with increased IgG4^+^ plasma cells (> 30 IgG4^+^ plasma cells/high power field, IgG4 to IgG ratio: around 50%) | - | - | - | - |
| IgG4 RLD #14 | - | M | IgG4-RLD | cholangitis and chronic pancreatitis | - | - | - | Prs p.o. |
| IgG4 RLD #15 | - | F | IgG4-RLD | vocal cord surgery, dense lymphoplasmacellular infiltrate, storiform fibrosis and accompanying phlebitis with marked infiltration of IgG4^+^ plasma cells | - | - | - | - |
| IgG4 RLD #16 | - | M | IgG4-RLD | recurrent infections mainly of the upper respiratory tract, depression, Mikulicz´s disease, swelling of the parotid gland and eyes, elevated IgG4 levels | - | - | - | naproxen p.o. 1000 mg/d, Prs p.o. |
